# Olfactory Bulb Volume and Asymmetry as Predictors of Executive Dysfunction in Adolescents with Congenital Heart Disease

**DOI:** 10.1101/2024.09.24.24314159

**Authors:** Adriana Racki, Anushka Shah, Ruby Slabicki, Julia Wallace, Vince K. Lee, Rafael Ceschin

## Abstract

**Background and Purpose:** Common sequelae for patients with congenital heart disease (CHD) are neurodevelopmental disabilities including executive function, attention, and socio-emotional deficits. Although these are common diagnoses for patients with CHD, limited research has investigated the mechanistic underpinnings of these findings. Our previous research examined the association between abnormal respiratory ciliary motion and brain abnormalities in infants with CHD. Results suggested that abnormal ciliary motion correlated to a spectrum of subtle dysplasia, notably within the olfactory bulb (OB)^1^. Our current study investigates whether OB anomalies predict neurodevelopmental outcomes for pediatric patients with CHD. We hypothesize that adolescents with CHD who exhibit aberrant morphological measurements in the OB are more likely to suffer from executive functional deficits.

**Materials and Methods:** A prospective, observational study of 54 CHD and 75 healthy subjects, ages 6-25 years old, was completed under the supervision of a senior pediatric neuroradiologist. T2 3D Space and T2 Blade 2MM MRI images were manually segmented to extract volumetric bilateral regions of the OB and cerebrospinal fluid (CSF) using ITK-SNAP. Imaging metrics were correlated to OB asymmetry, CSF to OB ratio, total CSF volume, total OB volume, and independent left and right CSF and OB volumes. Linear regression was used to evaluate MRI morphologic measurements with co-variates: CHD status, sex, MRI age, and segmenter. Executive function was determined by the Behavioral Rating Inventory of Executive Function (BRIEF) Parent Report and Delis-Kaplan Executive Function System (D-KEFS) for subjects ages 6-16. Cognition and olfactory function were measured with the NIH Toolbox Cognitive Battery and Odor Identification Test, respectively.

**Results:** No statistically significant results were reported between cohorts for asymmetry of OB, CSF to OB ratio, total CSF volume, total OB volume, nor between independent left and right CSF and OB volumes. Increased OB volume was associated with worse outcomes on the BRIEF Parent Report (p≤0.03). Asymmetry of OB predicted poorer executive functioning as reported by parents on the BRIEF (p≤0.05). Overall, the CHD cohort demonstrated worse scores on the BRIEF Parent Report compared to controls. Across groups, no significant association was reported for olfaction function measured by the NIH Toolbox Odor Identification Test on a limited subset of participants.

**Conclusion:** As survival rates for CHD improve, there is an increased risk of long-term neurodevelopmental impairments. Our findings identify adolescents who are at risk for executive dysfunction, particularly those showing increased OB volume and/or asymmetry of the OB. This is particularly concerning for the CHD population with atypical OB morphology, who also exhibit significantly poorer outcomes on the BRIEF Parent Report and face a higher overall risk. Increased OB volume and OB asymmetry are olfactory-based biomarkers that may help identify at-risk CHD patients earlier, enabling more timely intervention and support.

## INTRODUCTION

With enhanced surgical outcomes, we have seen a remarkable increase in life expectancy for individuals with congenital heart disease (CHD).^2^ This has translated into a higher prevalence of adults experiencing long-term neurodevelopmental disabilities including executive function, attention, and socio-emotional deficits. Though decades of research highlight the cognitive, learning and socio-behavioral deficits faced by individuals with CHD, our management of these patients remains inadequate.

Previous research suggests that cilia signaling plays a critical role in the pathogenesis of CHD with several identified CHD mutations arising from genes-related to cilia^3^. We know that disruptions in primary cilia impact embryonic development and left-right patterning of the heart contributing to the various cardiac lesions commonly seen in this population^4^. Consequently, patients with CHD exhibit a higher prevalence of abnormalities in motile cilia leading to an accumulation in extra-axial cerebrospinal fluid (CSF), delayed brain maturation, and subtle dysplasia within multiple anatomical regions of the brain, including the olfactory bulb (OB)^1^. These ciliary perturbations may be associated with the maladaptive neurodevelopmental outcomes observed in the CHD population, though limited research has explored this clinical interplay.

The primary aim of the study was to assess whether OB anomalies are prognostic of neurodevelopmental outcomes for adolescents with CHD by correlating morphological measurements of the OB to executive function via the NIH Toolbox Cognitive Battery, Behavioral Rating Inventory of Executive Function Parent Report (BRIEF) and Delis-Kaplan Executive Function System (D-KEFS) assessments. The asymmetry and aberrant shape of the OB were visualized using ITK-SNAP program in which volumetric measurements were correlated to 3D segmentation models.

## MATERIALS AND METHODS

### Participants

CHD and healthy participants were recruited prospectively from a single site academic center using integrated advertisements and the online registry, Pitt+Me. Participants with CHD were referred from outpatient cardiology clinics as well. All participants were native-English speakers ages 6 or older. Exclusion criteria for our study included cardiac transplantation, genetic syndromes, bleeding or neurological disorders, pacemaker or defibrillator status, pregnancy, and metal implants. All participants were excluded from our study analysis for any MRI contraindications. Healthy controls with a history of preterm birth, <34.0 weeks were excluded from the study. We initially screened 143 patients with CHD and 98 healthy controls (**Supplemental Figure 1**). A total of 69 CHD and 92 healthy controls underwent brain MRI scanning at 6-25 years old. After removing cases without analyzable T2-weighted images due to motion artifact, poor tissue contrast and image quality, the final sample of patients included the following: CHD patients (n=54) and age-matched controls (n=75), with ages ranging from 6-25 years. We have previously published work on this recruitment cohort^5–9^.These patients were prospectively recruited at our institution with Institutional Review Board approval and oversight (for reference: Institutional Review Board STUDY20060128 and STUDY1904003). The project was completed in accordance with the ethical principles of the Helsinki Declaration.

ITK-SNAP was used for image visualization and manual segmentation of anatomical regions of interest^10^. T2 3D Space and T2 Blade 2MM MRI images, formatted as NIFTI files, were uploaded into ITK-SNAP. The main program window was used with three orthogonal image views showing the axial, coronal and sagittal planes. Image contrast adjustments were made to optimize grey and white matter for accuracy of segmentation and consideration of acquisition parameters. **Figure 1** shows the general workflow for manual segmentation. Identification of the olfactory sulcus and groove were used to locate the OB. Linked crosshairs were placed on the OB to confirm the region of interest in all three planes. Using the sagittal plane, the OB and surrounding CSF were visualized and segmented from the lateral to medial margins. The coronal view was used to assess the anterior and posterior boundaries with correction for partial volumes. The axial view was also utilized to correct for partial volume averaging. Using an additional window view, a 3D surface rendering was generated based on manual segmentation results. Three segmenters analyzed the dataset, with overlap sets of subjects being segmented by multiple users to ensure segmentation consistency. A calibration dataset of 12 subjects was used where all segmenters independently segmented each structure, and a DICE coefficient was used to measure the degree of overlap. A minimum DICE score of 0.7 was required between all structures segmented by multiple segmenters. Structures with disagreement amongst segmenters (DICE < 0.7) were adjudicated under the guidance of a senior pediatric radiologist, who also inspected all final segmentations.

**Figure 1.**
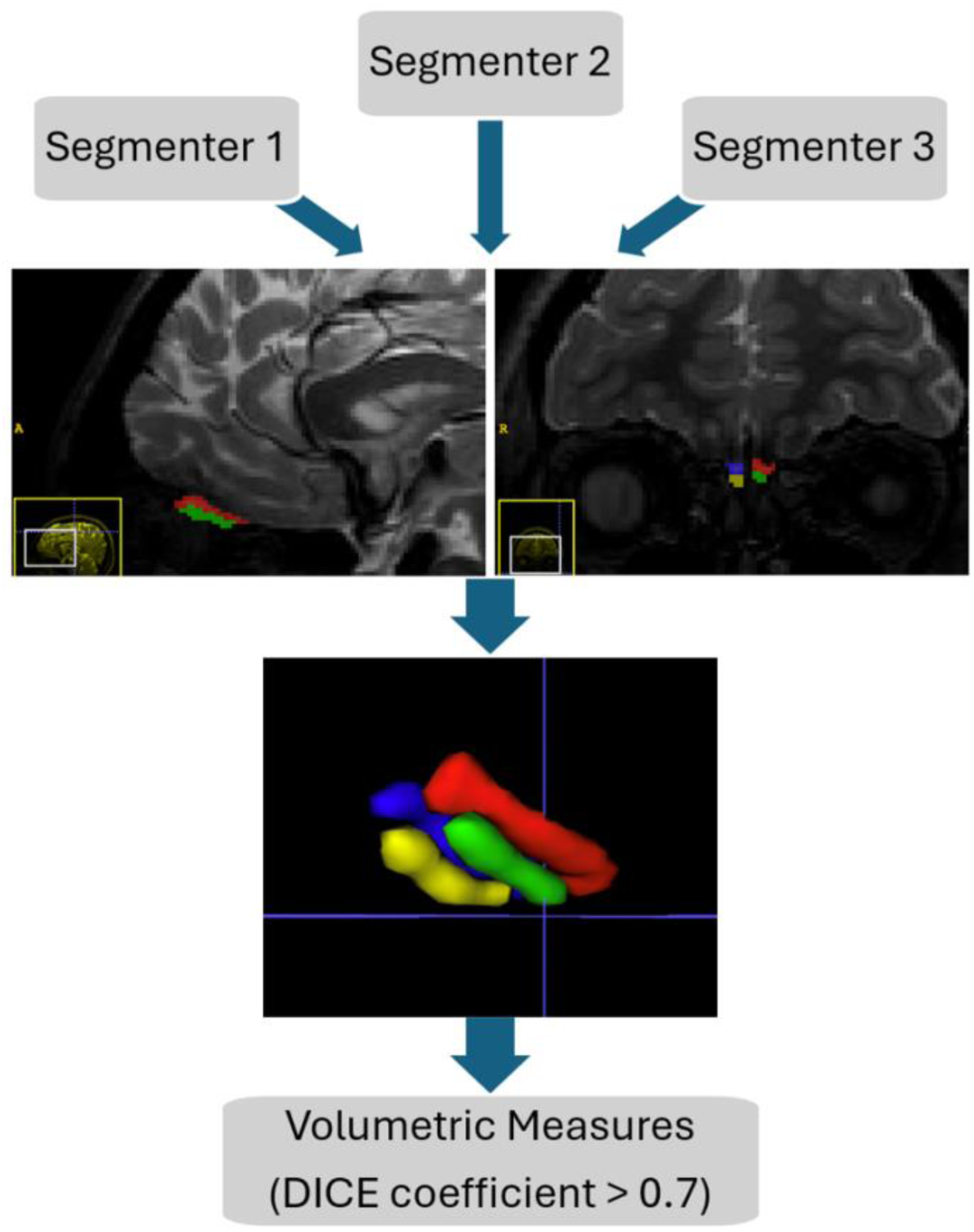
Olfaction Segmentation Flow Diagram ITK-SNAP software application was used for manual segmentation. Volumetric measurements were reported based on 3D segmentation results. Segmenters 1-3 independently completed manual segmentation of bilateral regions of the OB and CSF with 12 overlapping cases. Inter-rater reliability was assessed using a DICE score > 0.7. No significant difference was identified between inter-raters’ manual segmentation results. A senior pediatric neuroradiology oversaw manual segmentation of the OB and CSF with final approval of results. Abbreviations: OB = olfactory bulb. CSF = cerebrospinal fluid.

Group-wise differences were measured using Student’s t-test. We used linear regression to evaluate the association between neurocognitive outcomes and OB metrics, including CHD status, age, sex, and segmenter as covariates.

## RESULTS

### Participant Demographics

54 individuals with CHD were segmented out of the 143 CHD initially screened (**Supplemental Figure 1**). 75 healthy subjects were segmented out of 98 patients initially screened. There was a total of 59.26% of males segmented in the CHD group compared to 42.67% of males in the control group (**Supplemental Figure 2**). Across both cohorts the mean age at MRI scan was 14.40 years.

### Inter-rater Reliability

Three independent segmenters manually segmented the OB and CSF. There were 38 cases completed by 2 segmenters, with each case being evaluated by both segmenters (**Supplemental Figure 2**). To increase the reliability of the study, a DICE coefficient of greater than 0.7 was maintained. Additionally, across all regions of interest no significant difference was identified between inter-rater manual segmentation results.

### OB Metrics

Across cohorts, no statistical difference was seen for sex or age at MRI scan for asymmetry of OB, CSF to OB ratio, total CSF volume, total OB volume, nor between independent left and right CSF and OB volumes (**Figure 2**). No statistically significant result was reported between cohorts for asymmetry of OB, CSF to OB ratio, total CSF volume, total OB volume, nor between independent left and right CSF and OB volumes.

**Figure 2.**
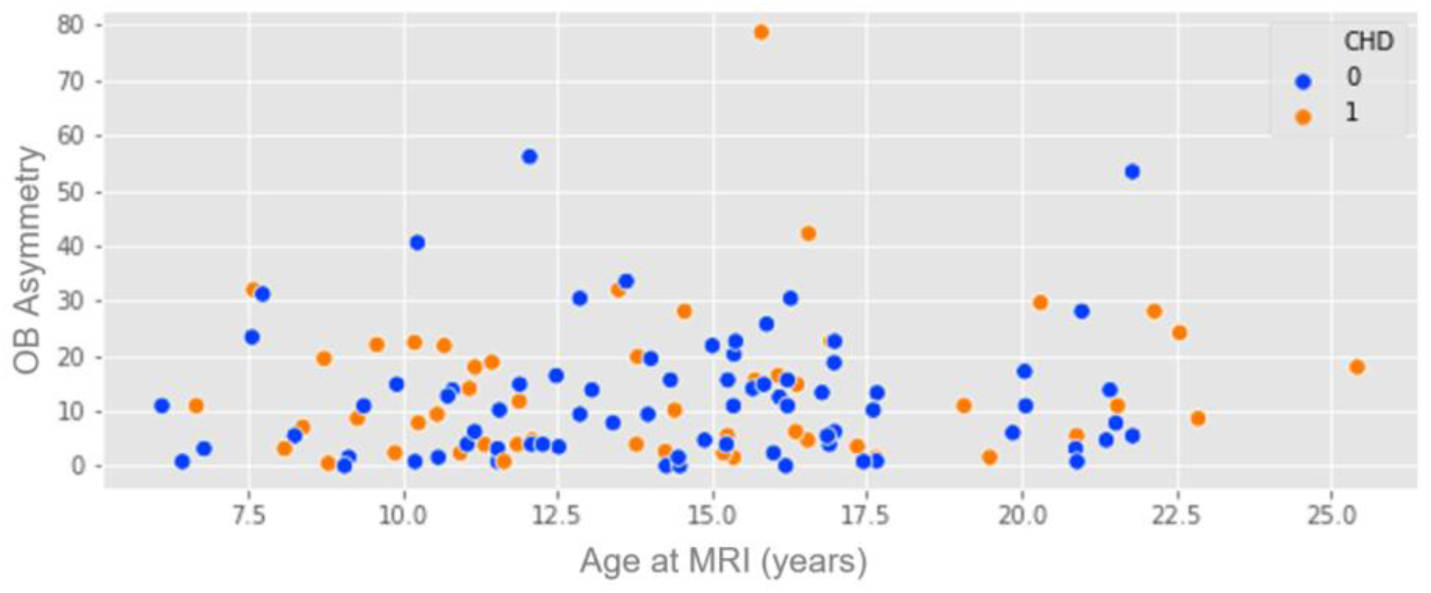
MRI Years versus OB Asymmetry [Blue= Control, Orange= CHD] Across cohorts, no statistically significant result was reported for asymmetry of OB based on age at MRI scan. Abbreviations: CHD = congenital heart disease. OB = olfactory bulb.

### Total OB Volume

Across both cohorts, an increased total OB volume predicted a worse score on the BRIEF Parent Report within the domains of Initiation (p≤0.03), Working Memory (p≤0.02), Global Executive Composite (p≤0.009), and Metacognition Index (p≤0.03) (**Table 1**, **Figure 3B**).

**Figure 3.**
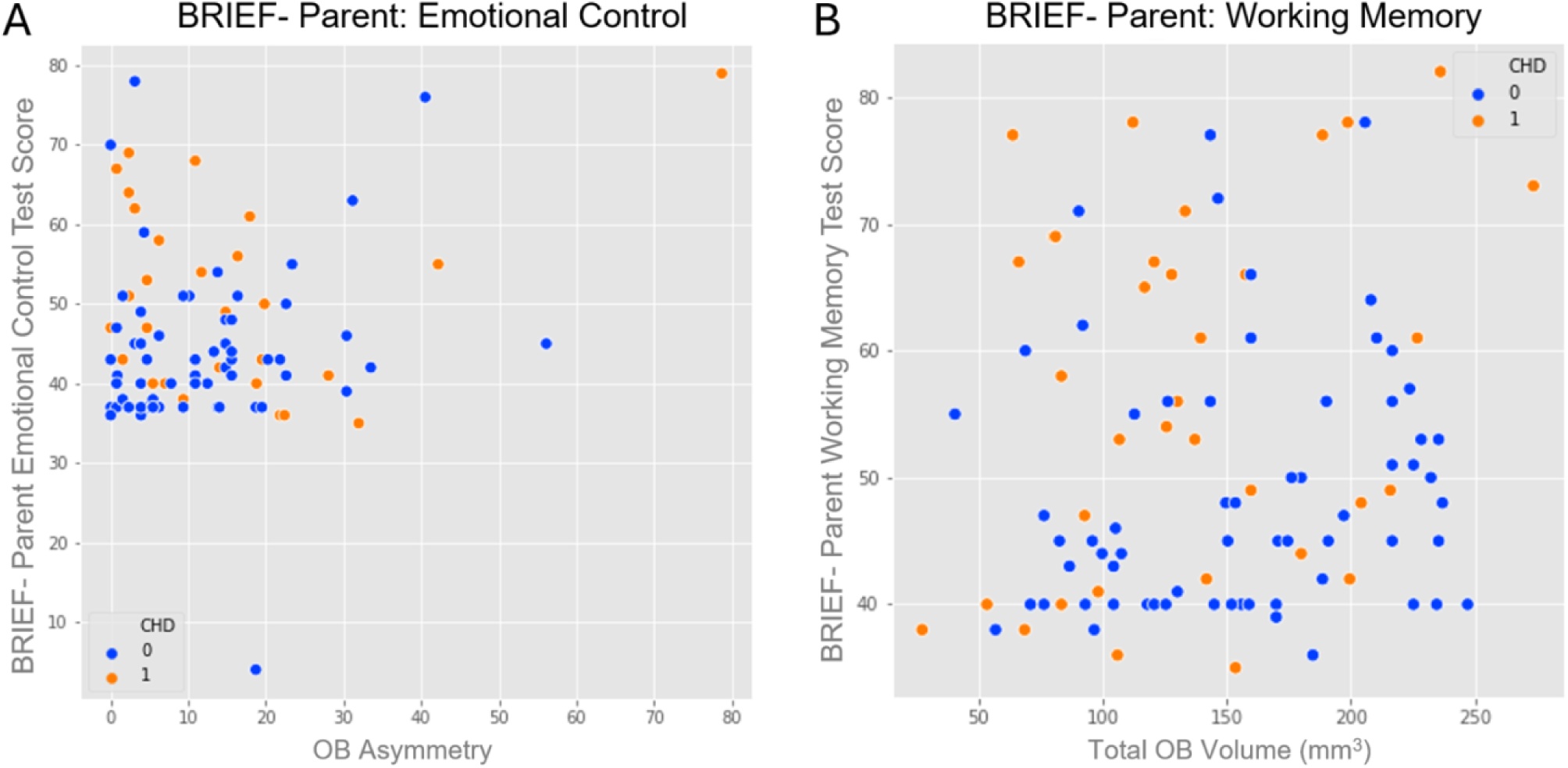
A and B. BRIEF-P Emotional Control and BRIEF-P Working Memory [Blue= Control, Orange= CHD] The CHD cohort demonstrated the worst performance on the BRIEF compared to controls (*p* ≤ 0.05). **A.** CHD and healthy controls with increased asymmetry of OB displayed poorer BRIEF Parent Report subdomain outcomes for Emotional Control (p≤0.03). **B.** Across both cohorts, increased total OB volume predicted worse score on the BRIEF Parent Report within the subdomain of Working Memory (p≤0.02). Abbreviations: CHD = congenital heart disease. OB = olfactory bulb.

**Table 1.** OB metrics and Neurocognitive Testing Regression Adolescents and their parents completed neurocognitive testing to assess subdomains of executive functioning. Morphological measurements of the OB were correlated to results from the NIH Toolbox Cognitive Battery, BRIEF Parent Report and D-KEFS assessment scores to assess neurodevelopmental outcomes. For CHD and healthy controls, an increased total OB volume predicted a worse score on the BRIEF Parent Report within the domains of Initiation (p≤0.03), Working Memory (p≤0.02), Global Executive Composite (p≤0.009), and Metacognition Index (p≤0.03). Increased asymmetry of OB displayed poorer BRIEF Parent Report subdomain outcomes for Inhibition (p≤0.03), Shift (p≤0.02), Emotional Control (p≤0.03), Planning & Organization (p≤0.05), Global Executive Composite (p≤0.03), Behavioral Regulation Index (p≤0.01), and Metacognition Index (p≤0.04). No significant association was reported for olfaction function for NIH Toolbox Odor Identification scores. Abbreviations: OB = olfactory bulb. BRIEF = Behavioral Rating Inventory of Executive Function. D-KEFS = Delis-Kaplan Executive Function System

| Neurocognitive Test | OB Asymmetry |  |  |  | Total OB Volume |  |  |  |
| --- | --- | --- | --- | --- | --- | --- | --- | --- |
|  | Coef | Std err | t | p | Coef | Std err | t | p |
| BRIEF- Parent: Initiation | 0.23 | 0.08 | 3.03 | <0.01 | 0.05 | 0.02 | 2.25 | 0.03 |
| BRIEF- Parent: Emotional Control | 0.17 | 0.08 | 2.25 | 0.03 | 0.03 | 0.02 | 1.57 | 0.12 |
| BRIEF- Parent: Working Memory | 0.04 | 0.08 | 0.50 | 0.62 | 0.05 | 0.02 | 2.43 | 0.02 |
| BRIEF- Parent: Global Executive Composite | 0.19 | 0.08 | 2.27 | 0.03 | 0.06 | 0.02 | 2.68 | 0.009 |
| BRIEF- Parent: Metacognition Index | 0.17 | 0.08 | 2.08 | 0.04 | 0.05 | 0.02 | 2.23 | 0.03 |
| BRIEF- Parent: Behavioral Regulation Index | 0.17 | 0.07 | 2.53 | 0.01 | 0.03 | 0.02 | 1.64 | 0.10 |
| BRIEF- Parent: Inhibition | 0.13 | 0.06 | 2.23 | 0.03 | 0.03 | 0.02 | 1.59 | 0.12 |
| BRIEF- Parent: Shifting | 0.17 | 0.07 | 2.48 | 0.02 | 0.04 | 0.02 | 1.93 | 0.06 |
| BRIEF- Parent: Planning | 0.16 | 0.08 | 2.00 | 0.05 | 0.04 | 0.02 | 1.94 | 0.06 |
| D-KEFS: Tower Test | 0.07 | 0.03 | 2.28 | 0.04 | -0.02 | 0.02 | -0.86 | 0.41 |
| D-KEFS: Trails Letter Sequencing | -0.03 | 0.02 | -1.93 | 0.06 | -0.004 | 0.004 | -0.88 | 0.38 |
| D-KEFS: Trails Letter Switching | -0.006 | 0.01 | -0.46 | 0.65 | 0.0009 | 0.004 | 0.25 | 0.81 |
| NIH Toolbox: Odor Identification Test | 0.09 | 0.12 | 0.73 | 0.47 | 0.04 | 0.03 | 1.34 | 0.18 |
| NIH Toolbox: Fluid Composite | -0.12 | 0.17 | -0.72 | 0.48 | 0.03 | 0.04 | 0.69 | 0.49 |
| NIH Toolbox: Crystallized Composite | -0.02 | 0.12 | -0.18 | 0.86 | 0.02 | 0.03 | 0.67 | 0.51 |

### Asymmetry of OB

CHD and healthy controls with increased asymmetry of OB displayed poorer BRIEF Parent Report subdomain outcomes for Inhibition (p≤0.03), Shifting (p≤0.02), Emotional Control (p≤0.03), Planning & Organization (p≤0.05), Global Executive Composite (p≤0.03), Behavioral Regulation Index (p≤0.01), and Metacognition Index (p≤0.04) (**Table 1**, **Figure 3A**). Across cohorts, asymmetry of OB demonstrated improved performance on the D-KEFS Tower Test (p≤0.04), which measures spatial planning, self-regulation, impulsivity, and problem-solving (**Table 1**).

### Cognitive Ability/Executive Function

Overall, the CHD cohort showed worse Fluid (p≤0.04) and Crystallized (p≤0.001) composite scores compared to the control group on the NIH Toolbox Cognitive Battery. Furthermore, the CHD cohort demonstrated worse performance on the BRIEF Parent Report across all executive domains assessed (**Figure 3A and B**). The CHD group performed worse on the D-KEFS Trails Letter Sequencing (p≤0.001) and D-KEFS Trail Letter Switching (p≤0.001).

### Olfaction Function

Due to the timing of data acquisition and subject availability, only 50 subjects completed the NIH Toolbox Odor Identification. Across both CHD and healthy control groups, no significant association was reported for olfaction function for NIH Toolbox Odor Identification scores (**Table 1**).

## DISCUSSION

### Increased Total OB Volume

Our study identified that increased OB volumes are associated with worse executive function. This result may be explained by the relationship of OB volume and pubertal timing^11^. During puberty, the size and function of the OB accelerates, starting in pre-puberty and following a consistent pattern with age^11^. Interestingly, there is strong evidence linking increased OB volume and precocious puberty^11^. This connection is further supported by the shared embryonic origin of the OB and gonadotropin-releasing hormone from the olfactory placode, as well as their combined role in the reproductive system^11^. Since the OB regulates behavior in response to chemical signals related to pubertal development and reproduction^11^, it is possible that the increased OB volume observed in our adolescent population may be reflecting this developmental pattern. More generally, variability in OB volumes has been shown in many acquired disorders including chronic rhinosinusitis, neurodegenerative disease, and congenital disorders^11^.

Alternatively, there have been two case reports of asymptomatic enlargement of the OB in Neurofibromatosis type-1^12^. Of these cases, one patient exhibited transient enlargement of the OB with a return to baseline size one year later^12^. This result may represent reversible microstructural alterations of the OB white matter, though it is unclear^12^. This observation is consistent with previous research, suggesting that microstructural changes, such as inadequate myelination, are associated with executive impairments in adolescents with CHD^13^. These functional impairments are likely attributable to a range of factors including alterations in gray matter volumes, connectivity and various social and environmental influences^13^. Our study results may represent this observation, as individuals with increased OB volume displayed a notable deficit on the BRIEF Parent Report, with dysfunction in Initiation, Working Memory, Global Executive Cognition, and Metacognition Index scales (**Table 1**). Our results highlight a significant concern for survivors of CHD, who showed markedly poorer outcomes on the BRIEF Parent Report compared to healthy controls (**Figure 3B**). This multifactorial process may be contributing to the executive dysfunction observed. Since our dataset does not follow patients longitudinally, it is unclear whether we are capturing a transient or more stable process. However, considering the broad age range of participants, 6-25 years, it is unlikely that our results represent a transient phenomenon.

### Asymmetry of OB

During our lifespan, we require asymmetrical organization within our hemispheres to properly tend to higher level processing, including spatial planning, executive functioning, and coordination of multi-task functions^14^. With aging, we see a progressive asymmetrical loss with prophesized thinning of hemispheric regions of the brain^15^, though limited studies have investigated the rate at which specific anatomical regions of the brain undergo thinning and asymmetrical atrophy. Notably, within our adolescent study population we see increased asymmetry of the OB (**Figure 4**). Limitations of our study methodology inhibit our ability to discern whether this asymmetric change is related to a nonproportionate shift in right versus left OB changes or related to thinning of the bulb. Since our participants are not longitudinally followed, we cannot confidently suggest whether this asymmetry represents early deviations from normal developmental patterns.

**Figure 4.**
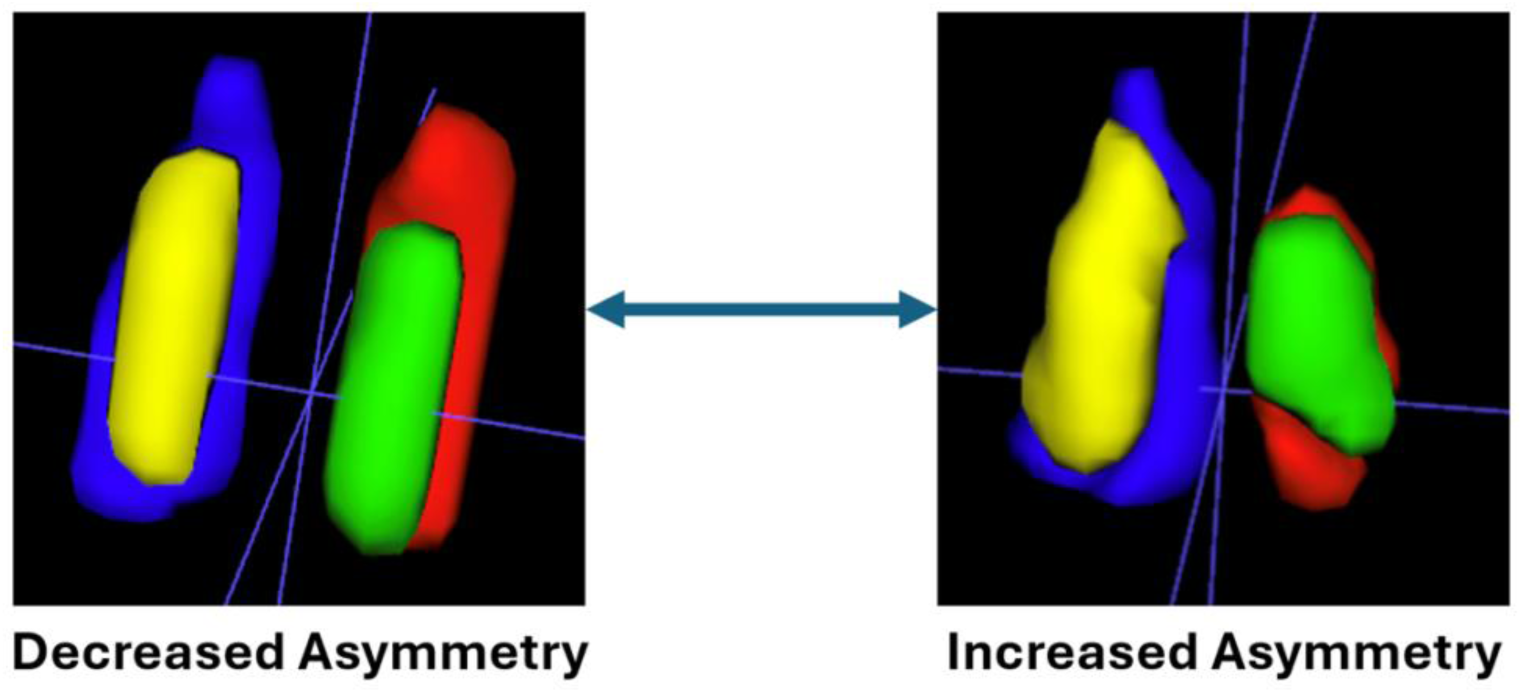
Decreased Asymmetry versus Increased Asymmetry ITK-SNAP application software after manual segmentation and 3D renderings depicts the OB morphology on a spectrum from decreased asymmetry to increased asymmetry. Across cohorts, asymmetry of OB predicted poorer executive functioning using BRIEF (*p* ≤ .05). Abbreviations: OB = olfactory bulb. BRIEF = Behavioral Rating Inventory of Executive Function.

Our findings indicate that adolescents with asymmetry in the OB perform worse on the BRIEF Parent Report, suggesting impaired executive functioning. Specific deficits were seen within the following subdomains: Inhibition, Cognitive Flexibility, Emotional Control, Planning & Problem Solving, Behavioral Regulation Index, Metacognition Index scales and Global Executive Composite. (**Table 1**). CHD patients exhibiting increased asymmetry represent a subgroup at highest risk of executive dysfunction, as these patients show the worst performance overall on the BRIEF Parent Report as compared to controls (**Figure 3A**).

Given the heterogeneity of the OB and limited research investigating OB structure and function, it is unclear whether this global executive deficit is related to asymmetry of the OB. However, prior research suggests that asymmetry in the OB may indicate a congenital disorder associated with genes involved in OB development^11, 16, 17^. Functional neuroimaging research has helped us understand that executive dysfunction involves a broad network of brain regions, extending beyond the prefrontal cortex to include the parietal cortex, basal ganglia, thalamus, among other areas^18^. Given that executive functioning is highly sensitive to disruptions,^18^ it is possible that OB asymmetry is affecting the executive function neural network, potentially contributing to the cognitive deficits observed in our population.

We know from research in children with attention-deficit hyperactivity disorder that underlying symptoms of inattention and impulsivity may be related to cerebral asymmetry among other apparent lateralized differences in the brain^19^. With further suggestion that asymmetry may also contribute to the progressive pattern of executive dysfunction seen in some children^19^. Importantly, we must consider the possibility that some children may not exhibit early neurodevelopmental influences, which might only become apparent after brain maturation^19^. This highlights the need to address both delayed development and the heterogeneity within our CHD population^19^.

On the contrary, asymmetry of OB demonstrated improved D-KEFS Tower Test scores, which incorporates spatial planning, self-regulation, impulsivity, and problem-solving abilities (**Table 1**). While statistically significant, the strength of this assertion cannot be adequately supported with an observation size of 17 participants. Given the limited research on OB structure and function, future studies may be helpful in better characterizing this result.

## LIMITATIONS

Limited research has explored the association between olfaction function and CHD. Studies show that odor identification increases throughout adolescence, peaking in the second decade of life^20^. Since olfactory identification seems to peak after the adolescence period^20^, limited literature exists in this age range. Some factors known to influence function include recurrent rhinosinusitis and aging, most specifically a decline in the fifth decade of life^20^. Past studies, in patients with autism spectrum disorder, attention-deficit hyperactivity disorder, and obsessive-compulsive disorder have shown olfactory impairment^21–23^, which were attributed to alterations in the central and peripheral olfactory systems^24^. Our findings suggest no significant difference in olfactory function, measured with the NIH Toolbox Odor Identification Test, across both adolescent groups (**Table 1**). Although, based on the timing of the data acquisition and availability of subjects, only 50 total participants completed the NIH Toolbox Odor Identification Test. Future research should further elucidate the association between olfactory function and adolescents with CHD.

As with all imaging techniques, partial volume averaging can influence the accuracy of manual segmentation of anatomical regions. We addressed this issue by employing inter-rater reliability measures with a threshold Dice coefficient > 0.7. Additionally, our study results were limited by the nature of our dataset, which was not longitudinal. Future studies should trend volumetric changes longitudinally to explain whether increased OB volume is a transient or stable process. Given the broad age range of our participants, it is unlikely that we are observing a temporary process. Future longitudinal studies may explore whether asymmetry is due to disproportionate changes between the right and left OB or related to thinning of the bulb.

With the ease of administration and brevity of the NIH Toolbox, it was selected to assess cognitive and olfactory function. While other cognitive assessments might offer greater specificity and sensitivity, administering a lengthier test could be challenging for our population, as evidenced by the fact that only 17 participants successfully completed the D-KEFS Tower Test. This underscores the suitability of the NIH Toolbox as the optimal choice for our study population.

## CONCLUSIONS

The notable improvement in survivorship of infants with CHD^25, 26^ has shifted our focus toward addressing long-term outcomes in brain function and neurodevelopment^27–29^. Here, we found that adolescents with increased OB volume and/or asymmetry of the OB demonstrated worse performance on the BRIEF Parent Report, depicting impaired executive functioning. These results display a significant concern for the CHD cohort, who demonstrated substantially worse outcomes on the BRIEF Parent Report compared to healthy controls. Our findings build on our past research, which identified subtle OB dysplasia in the CHD population,^1^ suggesting that OB dysmaturation may reflect functional deficits and serve as predictive biomarkers for neurodevelopmental outcomes in adolescents.

## Data Availability

Individual participant data that underlie the results reported in this article, after deidentification (text, tables, figures, and appendices), Study Protocol, and Statistical Analysis Plan, are available upon formal request. Requests should be submitted to the corresponding author.

## ACKNOWLEDGMENTS

I would like to extend my deepest gratitude to the PIRC staff and faculty for their invaluable support and dedication, which were crucial to the success of this project.

## ETHICS DECLARATIONS

The study describes the use of human data and involves human subjects; therefore, a decision from the Institutional Review Board for approval was required. All ethical guidelines have been followed, and necessary Institutional Review Board approvals have been obtained. Datasets were used only after formal Institutional Review Board approval. No simulated data or data openly available to the public was used before the initiation of the study. The Institutional Review Board of the University of Pittsburgh gave ethical approval for this work. The Institutional Review Board STUDY20060128: Multimodal Connectome Study approval 14 May 2014 and STUDY1904003 Ciliary Dysfunction, Brain Dysplasia, and Neurodevelopmental Outcome in Congenital approval 10 August 2016. The project was completed in accordance with the ethical principles of the Helsinki Declaration. Written informed consent was obtained from all participants, including consent from parents/guardians (including consent to publish), and the appropriate institutional forms have been archived. Any patient identifiers included were not known to anyone outside of the research group so cannot be used to identify individuals.

## Abbreviations

CHD: congenital heart disease

OB: olfactory bulb

CSF: cerebrospinal fluid

BRIEF: Behavioral Rating Inventory of Executive Function

D-KEFS: Delis-Kaplan Executive Function System

## SUPPLEMENTAL FIGURES

**Supplemental Figure 1.**
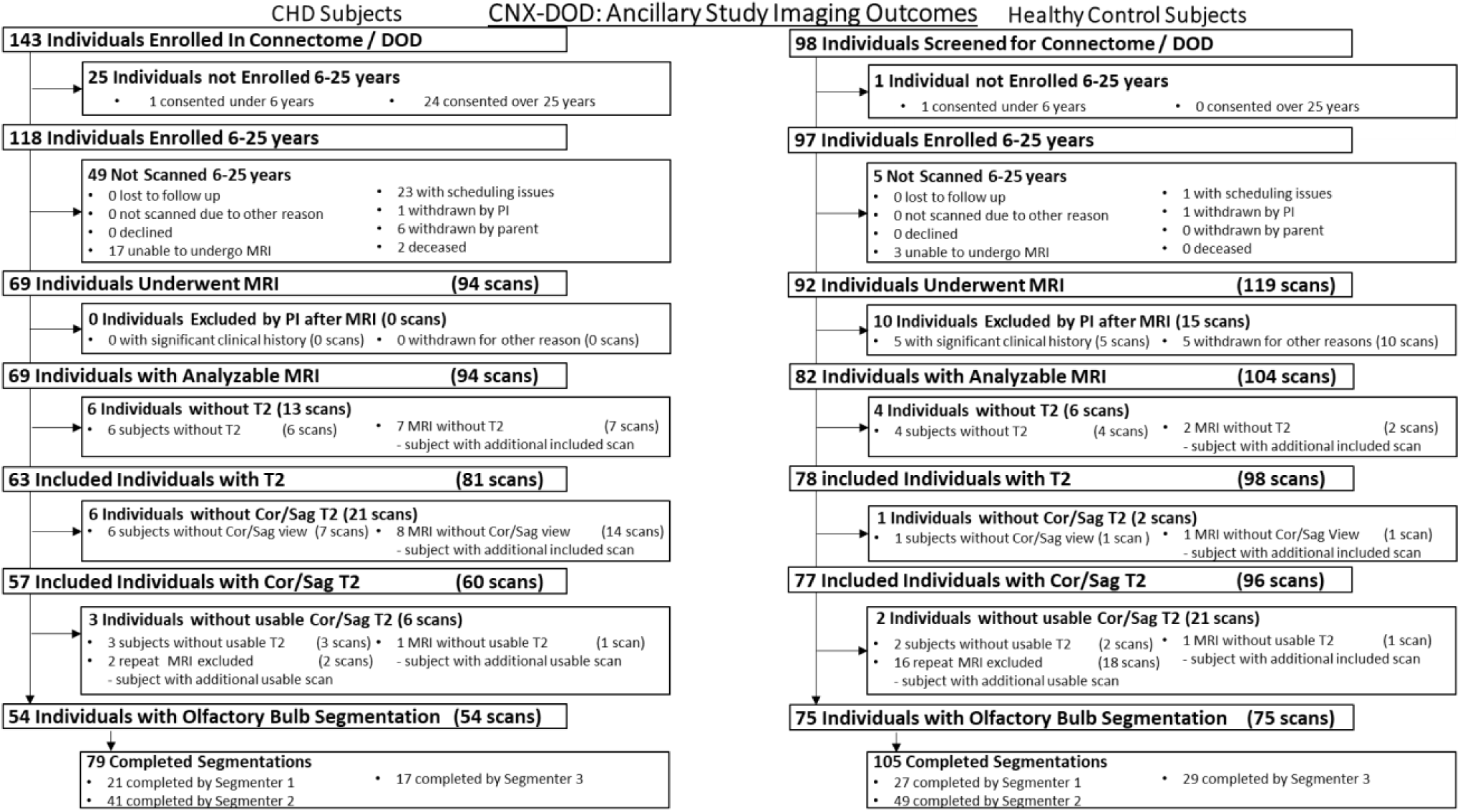
CNX-DOD: Recruitment Cohort and Exclusions A total of 69 CHD and 92 healthy controls underwent brain MRI scanning at 6-25 years old. Cases without analyzable T2-weighted images due to motion artifact, poor tissue contrast and image quality were removed from analysis. The final sample of patients included the following: CHD patients (n=54) and age-matched controls (n=75), with ages ranging from 6-25 years. The segmentation of all subjects were completed under the supervision of a senior pediatric neuroradiologist. Abbreviation: CHD = congenital heart disease

**Supplemental Figure 2.**
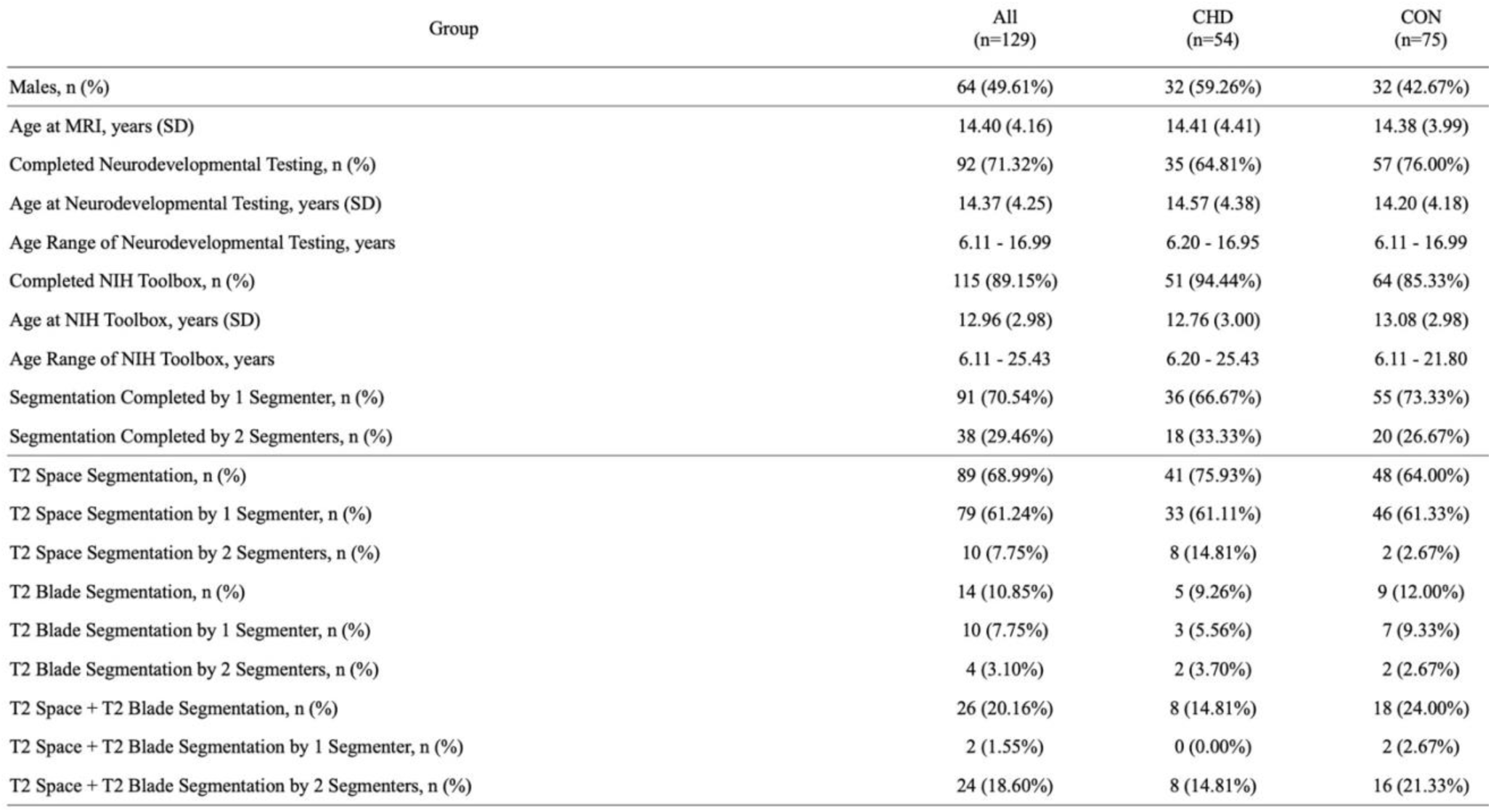
Cohort Demographics, Inter-rater reliability and MRI segmentations There was a total of 59.26% of males segmented in the CHD group compared to 42.67% of males in the control group. Across both cohorts the mean age at MRI scan was 14.40 years. There were 38 cases completed by 2 segmenters, with each case being evaluated by both segmenters. Abbreviation: CHD = congenital heart disease

